# In-kind nutritional supplementation for household contacts of persons with tuberculosis would be cost-effective for reducing tuberculosis incidence and mortality in India: a modeling study

**DOI:** 10.1101/2023.12.30.23300673

**Authors:** Pranay Sinha, Madolyn Dauphinais, Madeline E. Carwile, C. Robert Horsburgh, Nicolas A. Menzies

## Abstract

**Background:** Undernutrition is the leading cause of tuberculosis (TB) globally, but nutritional interventions are often considered cost prohibitive. The RATIONS study demonstrated that nutritional support provided to household contacts of persons with TB can reduce TB incidence. However, the long-term cost-effectiveness of this intervention is unclear.

**Methods:** We assessed the cost-effectiveness of a RATIONS-style intervention (daily 750 kcal dietary supplementation and multi-micronutrient tablet). Using a Markov state transition model we simulated TB incidence, treatment, and TB-attributable mortality among household contacts receiving the RATIONS intervention, as compared to no nutritional support. We calculated health outcomes (TB cases, TB deaths, and disability-adjusted life years [DALYs]) over the lifetime of intervention recipients and assessed costs from government and societal perspectives. We tested the robustness of results to parameter changes via deterministic and probabilistic sensitivity analysis.

**Findings:** Over two years, household contacts receiving the RATIONS intervention would experience 39% (95% uncertainty interval (UI): 23–52) fewer TB cases and 59% (95% UI: 44–69) fewer TB deaths. The intervention was estimated to avert 13,775 (95% UI: 9036–20,199) TB DALYs over the lifetime of the study cohort comprising 100,000 household contacts and was cost-effective from both government (incremental cost-effectiveness ratio: $229 per DALY averted [95% UI: 133–387]) and societal perspectives ($184 per DALY averted [95% UI: 83–344]). The results were most sensitive to the cost of the nutritional supplement.

**Interpretation:** Prompt nutritional support for household contacts of persons with TB disease would be cost-effective in reducing TB incidence and mortality in India.

**Summary:** Undernutrition is the leading cause of tuberculosis in India. Using a Markov state-transition model, we found that food baskets for household contacts of persons with tuberculosis would be cost-effective in reducing tuberculosis incidence and mortality in India.

**Research in context:** *Evidence before this study:* Undernutrition is the leading risk factor for TB worldwide. Recently, the RATIONS study demonstrated a roughly 40% reduction in incident TB among household contacts who received in-kind macronutrient and micronutrient supplementation.

*Added value of this study:* Although the RATIONS study demonstrated a dramatic reduction in incident TB, it is unclear if nutritional interventions to prevent TB are cost-effective. Previously, only one cost-effectiveness analysis of nutritional interventions for household contacts has been published. Due to lack of published data, that study had to make assumptions regarding the impact of nutritional interventions on TB incidence and mortality. In this study, we conducted an economic evaluation of a RATIONS-style intervention to reduce incident TB and mortality in India using observed data.

*Implications of all the available evidence:* In-kind nutritional supplementation for household contacts of individuals with TB disease would be cost-effective in reducing incident TB and TB mortality, particularly if TB programs leverage economies of scale to bring down the cost of the nutritional intervention.

## INTRODUCTION

Tuberculosis (TB) has regained its position as the leading infectious cause of death worldwide, with 10.6 million cases and 1.3 million deaths in 2022.(1) Undernutrition blunts the innate and adaptive immune response to *Mycobacterium tuberculosis*, the pathogen that causes TB.(2) At least 2.2 million of 10.6 million TB cases are attributable to undernutrition, making it the leading risk factor for TB.(1) There is also evidence that, among individuals diagnosed with TB disease, undernourished individuals face higher risks of death, treatment failure, and relapse despite undergoing antimycobacterial therapy.(2, 3) Historical data suggest that nutritional supplementation could have a beneficial role by reducing the incidence of TB disease and reducing case fatality among those with TB disease.(2) However, in the absence of contemporary empirical data, nutritional supplementation for persons with TB (PWTB) and their household contacts (HHCs) has not been widely adopted.

The recently published RATIONS study randomized more than 10,000 household contacts of persons with TB in eastern India between a control arm that received routine TB care with no additional in-kind nutritional supplementation and an intervention arm that received macronutrient and micronutrient support with a food basket that cost $13 per month for 6 months.(4) Compared to the control group, the intervention group experienced a 40% reduction in TB incidence over the following two years. PWTB in the RATIONS study, who also received nutritional support regardless of whether their household contacts were in the control or intervention groups, experienced an approximately 35% reduction in mortality compared to a previous National TB Elimination Program cohort. The RATIONS study shows the potential nutritional supplementation to prevent TB incidence and mortality among household contacts of PWTB.

In this study, we developed a Markov state transition model to explore if a RATIONS-style nutritional intervention would be cost-effective for reducing TB incidence and mortality in India.

## METHODS

### Study Design

We used a Markov cohort model to estimate the long-term costs and health outcomes of providing a food basket to HIV-negative household contacts of PWTB in India. We parameterized intervention costs and effect sizes from the results of the RATIONS study, which was conducted among household contacts of PWTB in Jharkhand, India, in a largely rural population with a high degree of multidimensional poverty.(4) We conducted the study from the perspective of India’s central government, which funds the National TB Elimination Program (NTEP). Recognizing that patient costs can often be substantially larger than provider costs, we also conducted a secondary analysis to estimate results from a societal perspective, considering costs borne by both patients and the Indian government. We programmed the model using TreeAge Pro v.22.2.0 (Tree Age, Williamstown, MA).

### Intervention strategies

We modeled two strategies. The “usual intervention” strategy reflects the status quo wherein household contacts and PWTB do not receive any in-kind nutritional support. In the RATIONS intervention, HHCs of PWTB receive a food basket that contains a daily multi-micronutrient tablet, rice, and lentils for 6 months.(4) The food basket provides 23g of protein and a total of 750 Kcal calories daily. In addition, under the RATIONS intervention we assumed that individuals progressing to TB disease would receive a nutritional supplement that provided 1200 Kcal with 52g of protein for all six months of TB treatment.

### Study Model

The Markov model simulates the natural course of TB incidence, cure, and sequelae for a starting cohort without TB disease. The model was constructed as a set of mutually exclusive health states, with natural history transitions represented by movements between these health states: development of TB disease, TB cure, death from TB, and death from other causes (Figure 1). Parameters defining TB incidence rates were specified to represent differences in the rates of progression to TB disease based on individual nutritional status. Individuals developing TB disease could die, self-cure, or be cured via treatment. Individuals who survive TB transition to an early post-treatment state, where they have an elevated risk of recurrent disease for 12 months as well as increased all-cause mortality. Following 12 months in the early-post treatment phase, individuals transition to the late post-treatment phase, where they continue to have increased all-cause mortality, but where their risk of TB disease falls to baseline. We assumed that after surviving TB, individuals would have a lower average quality of life due to chronic post-TB lung disease. We used the model to simulate progression of a cohort of 100,000 household contacts of PWTB through these health states over their lifetime, with a 6-month time step.

**Figure 1:**
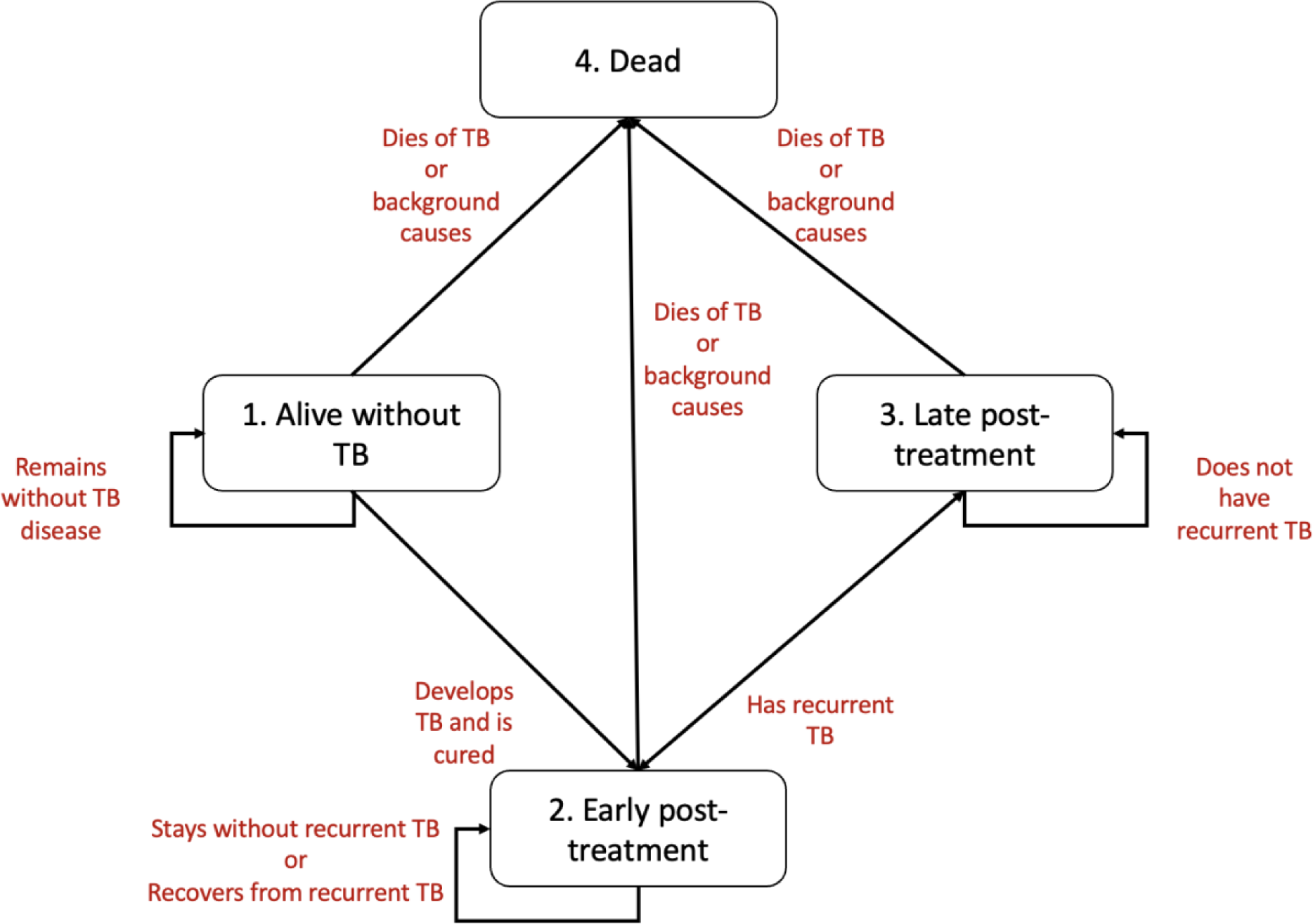
State transition diagram of model.

### Parameter inputs

#### Health Outcomes

Outcomes from the simulation included TB disease incidence, TB deaths, disability-adjusted life years (DALYs), and costs from the government perspective. In a secondary analysis, we also considered costs from a societal perspective. All outcomes were assessed over the lifetime of the modelled cohort.

We sourced TB incidence rates for HHCs after exposure from a previous meta-analysis.(5) Table 1 presents incidence estimates adopted for the analysis for the first five years. After five years of elevated risk of TB disease, we assumed the incidence would drop to prevailing rates in the general Indian population based on WHO estimates. (3) To account for the long-term secular declines in population TB incidence rates, we assumed a 3% annual decline based on incidence trends estimated for India over the past five years (Supplementary Figure 1). In the early post-treatment phase, we assumed that TB survivors had an increased risk of recurrent TB based on observed data from India.(6) In the late post-treatment phase, the risk dropped to baseline.

**Table 1:**
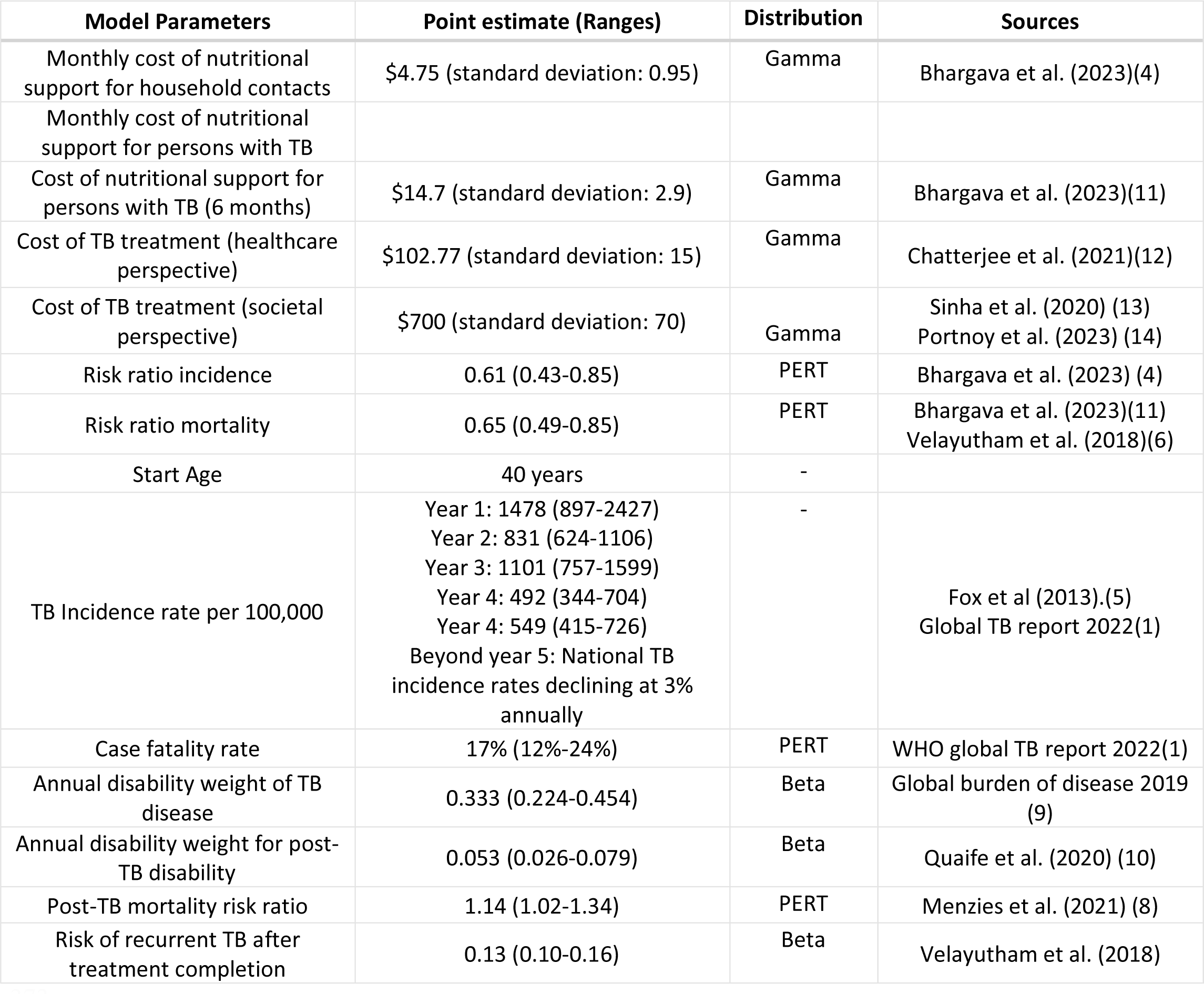
Model assumptions.

We obtained TB case fatality rates from the WHO Global TB Report 2022.(3) For background mortality rates, we constructed TB-deleted life tables using WHO life tables.(7) For individuals surviving TB, we estimated a lifetime increase in TB-attributable mortality using previous conservative estimates.(8)

To calculate DALYs, we obtained disability weights for TB disease from the Global Burden of Disease Study 2019.(9) We estimated the disability weight for TB survivors (due to persistent sequelae) to be 0.053 based on a published study of the global burden of post-TB lung disease.(10)

#### Intervention impact

We assumed that prompt nutritional supplementation would reduce the risk of incident TB disease among HHCs. Based on the results of the RATIONS study, we assumed an incidence rate ratio of 0·61 (95% CI 0·43, 0·85) for HHCs receiving supplementation compared to no supplementation for two years from the beginning of the intervention. After two years, we assumed that those receiving the intervention and those receiving usual care had a similar risk of incident TB disease. We assumed that PWTB who receive nutritional support have reduced TB-related mortality based on a comparison of outcomes of PWTB in the RATIONS study to an NTEP cohort, operationalized as a risk ratio of 0·65 (95% CI: 0·49, 0·85) applied between TB incidence and cure.(6, 11)

We used cost data from the RATIONS study, which reported supplementation costs of $13 per month inclusive of delivery costs. In sensitivity analyses, we examined a wide range of values for the cost of the nutritional supplementation, to account for regional variation, diversion of food grains, and wastage. We obtained TB treatment costs from a recent published study.(12) For costs borne by TB patients and affected households, we assumed that the direct medical costs, direct non-medical costs, and productivity losses borne by TB-affected household would be $700 (SD: 140) per TB episode.(13, 14) We rounded all costs to the nearest dollar. All costs are reported in 2022 US dollars.

### Cost-effectiveness analysis

We employed the model to compare the RATIONS nutritional supplementation intervention to the status quo. We projected TB cases, TB deaths, disability-adjusted life-years (DALYs) attributable to TB, and costs for each strategy over the lifetime of the study cohort. We calculated incremental cost-effectiveness ratios (ICERs) as the ratio of mean incremental costs to mean incremental health benefits, for the intervention compared to the usual care strategy. Costs and DALYs were discounted at 3% per annum.(15)

#### Willingness-to-pay

We derived our willingness-to-pay (WTP) threshold from Ochalek et al., who estimated country-specific willingness-to-pay thresholds based on the opportunity cost of healthcare spending.(16) For India, this study estimated a WTP threshold equivalent to 23% of per-capita GDP (US$550 in 2022). We considered ICERs below this threshold to be clearly cost-effective.

### Sensitivity analysis

To assess the robustness of our results, we conducted both deterministic and probabilistic sensitivity analyses.(17) For the deterministic sensitivity analysis, we defined *a priori* feasible ranges around core parameter values (Table 1) and then repeated the analysis multiple times, each time varying one parameter between the maximum and minimum of the feasible range, with other parameters held at their point estimate. Next, we conducted two-way sensitivity analyses, in which we simultaneously varied the parameters that emerged as the most important from one-way sensitivity analyses.

Lastly, we performed probabilistic sensitivity analysis (PSA). We defined uncertainty in parameter values as probability density functions (PDFs) for each model parameter. We then employed 2^nd^-order Monte Carlo simulation to repeat the analysis 10,000 times, each time drawing a random value for each model parameter from its PDF. This produced 10,000 estimates for each model outcomes, with the distribution of these estimates quantifying the uncertainty in model outcomes due to the combined uncertainty from all model parameters. We report the PSA results using cost-effectiveness acceptability curves.(18) In addition, we report uncertainty around main analysis results using 95% uncertainty intervals calculated from the PSA results.

## RESULTS

### Base Case Results

Base case results are presented in Table 2. Per 100,000 household contacts, those receiving the RATIONS intervention had lower cumulative TB incidence over their lifetime compared to those receiving usual care (6749 cases [95% uncertainty interval (UI): 5524–8520] vs. 7718 [95% UI: 6261– 9734]), risk ratio (RR) 0.87 [95% UI: 0.83–0.93]). In the first two years of this model, there were 1580 TB cases (95% UI: 1074–2461) in the RATIONS intervention as opposed to 2577 TB cases (95% UI: 1898–3634) with usual care, RR 0.61 (0.48–0.77). These results are consistent with the empirical findings of the RATIONS trial.

**Table 2:** Outcomes per 100,000 individuals. (DALYs: disability-adjusted life years; ICER: incremental cost effectiveness ratio [$/DALYs averted]. Costs and DALYs discounted)

| Strategy | DALYs (95% CI) | Healthcare cost (95% CI) | Healthcare ICER (95% CI) | Societal cost (95% CI) | Societal ICER (95% ICER) |
| --- | --- | --- | --- | --- | --- |
| Usual care | 41,172<br>(35,673-49,015) | \$0.76 million<br>(0.52-1.11) | Ref | \$5.24 million<br>(2.97-5.68) | Ref |
| RATIONS | 27,397<br>(22,208-35,452) | \$ 3.93 million<br>(2.89-5.18) | \$229 (133-387) | \$7.76 million<br>(5.71-10.58) | \$184<br>(83-344) |

The cohort receiving the RATIONS intervention was also estimated to have lower lifetime TB mortality (906 [95% UI: 667–1288] vs. 1529 [95% UI: 1196–2044] TB deaths, RR 0.59 [95% UI: 0.49–0.72]). In the first two years, there were 198 TB deaths (95% UI: 127–335) under the RATIONS intervention vs. 479 (95% UI: 343–710) under the usual care strategy (RR 0.41 [95% UI:0.31–0.56]). Per 100,000 individuals, the RATIONS intervention averted 13,775 DALYs (95% UI: 9036–20,199) compared to usual care. The majority of DALYs were averted in the RATIONS arm through reductions in TB mortality. Disaggregated DALY estimates for each strategy are reported in Table 3.

**Table 3:** Sources of DALYs per 100,000 in the two strategies.

| Component of Disability-adjusted Life Years | Usual care | RATIONS |
| --- | --- | --- |
| Years lost to TB mortality | 29,705<br>(22,820-40,615) | 16,951<br>(12,298-24,682) |
| Years lost to incremental post-TB mortality | 6,209<br>(2087-13,596) | 5,760<br>(1958-12,589) |
| Years lost to disability from TB disease | 1,245<br>(877-1784) | 1,071<br>(756-1538) |
| Years lost to disability from post-TB chronic lung disease | 4,013<br>(2910-5544) | 3,615<br>(2626-5027) |
| Total | 41,172<br>(35,673-49,015) | 27,397<br>(22,208-35,452) |

Under the governmental perspective total (discounted) costs for the usual care strategy were $0.76 million (95% UI: 0.52–1.11) per 100,000 individuals, representing the costs of treatment for individuals developing TB. Total governmental costs under the RATIONS intervention were $3.93 million (95% UI: 2.89–5.18), with $0.66 million (95% UI: 0.45–0.96) of this total due to the costs of TB care, and the remainder due to the costs of the nutritional intervention. Under the societal perspective total cost were $5.24 million (95% UI: 2.97–5.68) for the usual care strategy. Total societal costs under the RATIONS intervention were $7.76 million (95% UI: 5.71-10.58).

The incremental cost-effectiveness ratio of the RATIONS intervention, as compared to the status quo, was $229 per DALY averted (95% UI: 133–387) under the healthcare perspective and $184 (95% UI: 83– 344) under the societal perspective. Both these estimates are below the country specific WTP threshold of $550 per DALY averted.

### Sensitivity analysis

One-way sensitivity analyses for the general population showed that the two parameters with the greatest impact on the ICER were the pre-recipient cost of the RATIONS supplementation (baseline assumption: $28.5, range: $20-100) and the risk ratio of TB incidence for those receiving the nutritional supplementation (baseline assumption: 0.61, range: 0.43 to 0.85) (Table 4). In threshold analyses, the cost-effectiveness ratio for the RATIONS intervention was below to the WTP threshold ($550 per DALY averted) when the cost of supplementation was less than $75, indicating that the intervention would be cost-effective for any supplementation cost below this value. Even when we assumed the risk ratio for TB incidence was 0.99, the WTP threshold was not exceeded. The model was less sensitive to the risk reduction in TB mortality, the cost of TB treatment, and the disability weight for post-TB lung disease (Table 4).

**Table 4:** Results of one-way sensitivity analyses from a healthcare perspective.

| Variable Description | Variable Low | Variable Base | Variable High | Impact on ICER | ICER Low | ICER High |
| --- | --- | --- | --- | --- | --- | --- |
| Incremental cost of supplement for HHC (\$) | 20 | 28.5 | 100 | Increase | 125 | 675 |
| Risk ratio of TB incidence | 0.43 | 0.61 | 0.85 | Increase | 132 | 288 |
| Risk ratio of TB mortality | 0.49 | 0.65 | 0.85 | Increase | 147 | 262 |
| Case fatality rate in usual group | 0.12 | 0.17 | 0.24 | Decrease | 142 | 237 |
| Incremental cost of supplement for PWTB (\$) | 40 | 88.2 | 200 | Increase | 161 | 236 |
| Probability of TB recurrence | 0.04 | 0.06726 | 0.12 | Decrease | 171 | 191 |
| Cost of TB treatment (\$) | 100 | 700 | 300 | Decrease | 214 | 229 |
| Annual disability weight of post-TB lung disease | 0.0265 | 0.053 | 0.0795 | Decrease | 181 | 186 |
| Risk ratio of post-TB mortality | 1.02 | 1.14 | 1.34 | Decrease | 183 | 184 |

In two-way sensitivity analyses, the RATIONS intervention was cost-effective across a large range of costs of the supplement and risk ratio of TB incidence (Figure 3). When the cost of the supplement for HHCs was below $56, the intervention was always cost-effective from the government perspective. When considering the societal perspective, the intervention remained below the WTP threshold as long as the supplementation cost was below $59. We also conducted a two-way sensitivity analysis of the risk ratios for TB incidence and TB mortality. Even at the lowest estimates of the risk ratio for TB incidence (0.85) and TB mortality (0.85), the RATIONS intervention remained cost-effective with an ICER of $514.

**Figure 3:**
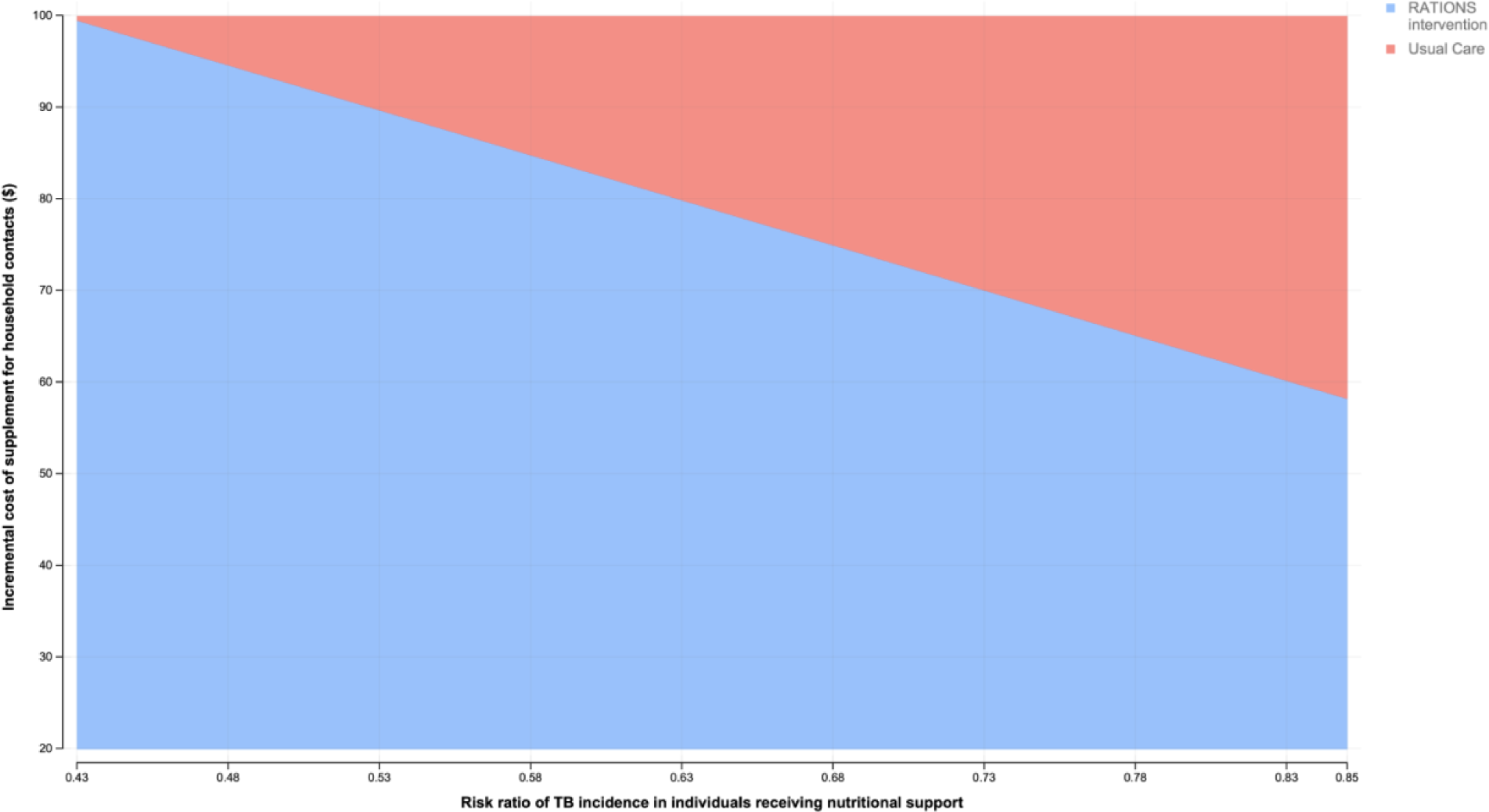
Two-way sensitivity analysis of the cost of intervention and risk ratio of TB incidence. Areas shaded blue denote that the intervention would be clearly cost-effective and red shaded regions indicate that the willingness-to-pay threshold of $550 was exceeded from the government perspective and usual care is preferred.

**Figure 4:**
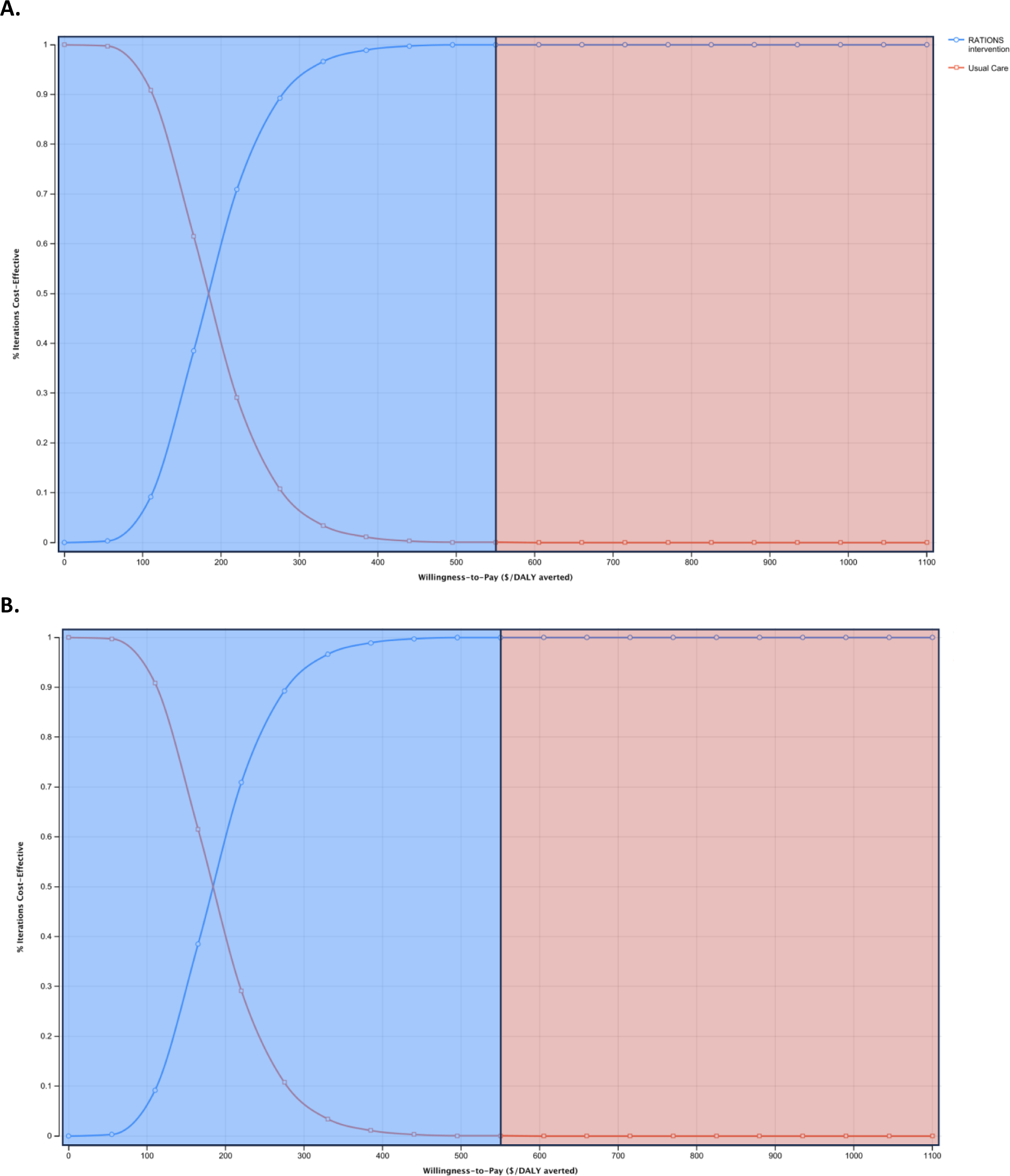
Cost-effectiveness acceptability curves. Cost-effectiveness acceptability curves for the models with A. government perspective and B. societal perspective. The blue line denotes the RATIONS intervention, and the red line indicates usual care. X-axis denotes willingness-to-pay and the Y-axis denotes the percentage of iterations (out of 10,000 iterations) in which a particular strategy was favored for a particular willingness-to-pay threshold. Areas shaded blue are below the willingness-to-pay threshold of $550 and those shaded red are above that threshold.

Lastly, in probabilistic sensitivity analysis, the RATIONS intervention was favored at a WTP of $550 in 99.95% and 99.97% of model simulations from both the government and societal perspectives, respectively. Indeed, RATIONS was consistently preferable in >99% of iterations at a WTP of $440. From a societal perspective RATIONS was consistently favorable in >99% of iterations at a WTP of $396. Cost effectiveness acceptability curves demonstrate the points at which RATIONS becomes consistently cost-effective (Figure 3).

## DISCUSSION

Nutritional interventions to mitigate risks of TB incidence and mortality are often perceived to be cost prohibitive. However, the RATIONS study has provided empirical evidence that low-cost nutritional support could have a substantial impact on lowering the risk of incident TB in settings with high rates of multidimensional poverty and undernutrition. Our analysis suggests that an in-kind nutritional intervention modelled on the food basket used in the RATIONS study could be cost-effective for reducing TB incidence and mortality in India. We estimated that RATIONS-style nutritional support would have a cost-effectiveness of $229 per DALY averted from a healthcare perspective and $184 per DALY averted from a societal perspective. Our findings can help national TB programs as they consider the economics of implementing nutritional support to reduce the incidence of TB among household contacts.

Balanced nutritional interventions likely reduce incident TB disease by enhancing the humoral and innate responses to *Mycobacterium tuberculosis*; these responses can help infected individuals eliminate or contain the bacilli.(2) Nutritional support to malnourished individuals may acutely restore their immune response. Indeed, mouse models suggest that providing a protein-rich diet to protein-deficient mice can restore their anti-tubercular immune response within two weeks.(19) Given that the majority of TB disease in high burden settings arises from infection within the past two years, nutritional support should be provided to HHCs promptly after the diagnosis of the index case to prevent incident TB disease.(20)

In our model, we limited the reduction in TB incidence among HHCs to two years after exposure since this was the duration of follow-up observed in the RATIONS study. For subsequent years, we assumed that individuals receiving the RATIONS intervention would have the same incidence rate as the usual care group, which remained elevated in years 3, 4, and 5 since exposure. Given the impact of nutritional support on the immune response to TB, it is possible that individuals receiving improved nutrition are more resistant to new TB infection. If so, the incidence reductions produced by the RATIONS intervention could extend beyond the two years assumed in this analysis. As such, our health impact and cost-effectiveness estimates may be conservative.

It is important to note that that the RATIONS study—on which this cost-effectiveness analysis was based—was conducted in a population with high levels of material deprivation and undernutrition, and TB incidence rates higher than those estimated for other parts of India. It is possible that the impact and cost-effectiveness of nutritional support may be lower in settings with a lower prevalence and severity of undernutrition, or lower TB incidence. Our two-way sensitivity analysis suggests that even at the lowest estimates of risk reduction for TB incidence (15% reduction) and mortality (15% reduction), the intervention would remain below the WTP threshold. These are credible effect sizes in less impoverished populations. A systematic review, which largely drew on data from high income countries, found a log-linear relationship between BMI and TB incidence wherein TB incidence decreases by approximately 14% per 1kg/m^2^ increase in BMI which is entirely achievable in 6 months on the RATIONS intervention. Further, a systematic review of randomized trials of macronutrient supplementation in PWTB, despite being underpowered, estimated that macronutrient supplementation was associated with a RR of 0.34 (95% CI: 0.10–1.20) for TB mortality.(21)

Public health investments to eliminate TB and undernutrition have historically provided some of the greatest returns on investment.(22, 23) The ICER estimated for nutritional support in our study is considerably below both the country-specific WTP threshold for India and the ICER for well-accepted TB preventive interventions, such as test-driven isoniazid preventive therapy for persons with HIV/AIDS ($1154 per DALY averted).(24) In our model, the ICER was most sensitive to the cost of nutritional supplementation for HHCs. Even when we assumed the lowest possible effect of nutritional interventions on incident TB disease, nutritional support would be cost-effective at $56 per recipient over 6 months, which was nearly twice the cost of the intervention. It is possible that state and national governments would enjoy economies of scale, not possible in the context of a clinical trial, that consequently could further lower the cost of the intervention.

Our analysis has several limitations. First, we did not model the impact of nutritional interventions on reducing early relapses and disengagement from care, even though these are plausible effects.(2, 25) Second, we did not account for reductions in TB transmission and secondary TB cases that might result from lower TB incidence, and instead focused on the direct health benefits experienced by intervention recipients. Third, our cost assumptions were limited to drug-susceptible TB, which is less costly to treat than drug-resistant TB, (13) and we also did not model the medical and non-medical costs faced by PWTB prior to their diagnosis. Approximately two-thirds of Indian PWTB treated in the public sector first seek care in the private sector and accrue large costs before an accurate TB diagnosis is made.(13) Forth, the model did not capture the positive externalities of nutritional support, including improvements in nutritional status and functional recovery and improved economic productivity among TB survivors, which have been explored in a previous analysis.(26) Taken together, these limitations suggest that our ICER estimates will be conservative, and that nutritional support could be even more cost-effective than we project.

Improvements in population-level nutrition and progress on social determinants of health likely contributed to the massive decline in TB incidence and mortality in countries like the United States and the United Kingdom that occurred even prior to the introduction of antimicrobials.(25) The striking findings of the RATIONS study have demonstrated the role nutritional interventions can play in the contemporary era to accelerate TB elimination. India is well positioned to institute a large-scale nutritional support program as it already has well-established infrastructure for delivering in-kind rations, mid-day meals, and nutritional counseling.(27-29) The TB Mitra Program, which crowdsources nutritional support for persons with TB, could also be scaled up.(30)

Additional research is necessary to help us understand how to target nutritional interventions equitably and efficiently. Operational studies are needed to design optimal delivery systems. Even as nutritional interventions are operationalized in settings with lower prevalence of undernutrition in India, epidemiological studies can help us understand the effect size of nutritional interventions in diverse populations. We also need clinical and translational studies to better define the nutritional needs of special populations, such as pregnant women and children, to develop the best possible nutritional package. Economic evaluations should be conducted alongside nutritional intervention studies to inform policy.

In conclusion, our analysis projected that a RATIONS-style intervention would be cost-effective for populations with a high dual burden of TB and undernutrition in India and drive large reductions in TB incidence and mortality among household contacts of persons with TB.

## FUNDING

This work was supported by the National Institutes of Health [grant number K01AI167733-01A1 to PS]; the Burroughs Wellcome/ASTMH Postdoctoral fellowship to PS; US Civilian Research and Development Foundation [grant number USB-31150-XX-13 to PS] with federal funds from the Government of India’s Department of Biotechnology, the Indian Council of Medical Research, the National Institutes of Health, the National Institute of Allergy and Infectious Diseases, and the Office of AIDS Research and distributed in part by CRDF Global [DAA3-19-65673-1]; grant from the Warren Alpert Foundation to PS); and a career investment award by the Department of Medicine at Boston University Chobanian & Avedisian School of Medicine. The funders had no role in study design, analysis, or reporting.

## CONFLICTS OF INTEREST

We do not report any conflicts of interest.

## Data Availability

All data produced in the present study are available upon reasonable request to the authors

---

**Supplementary Figure 1:**
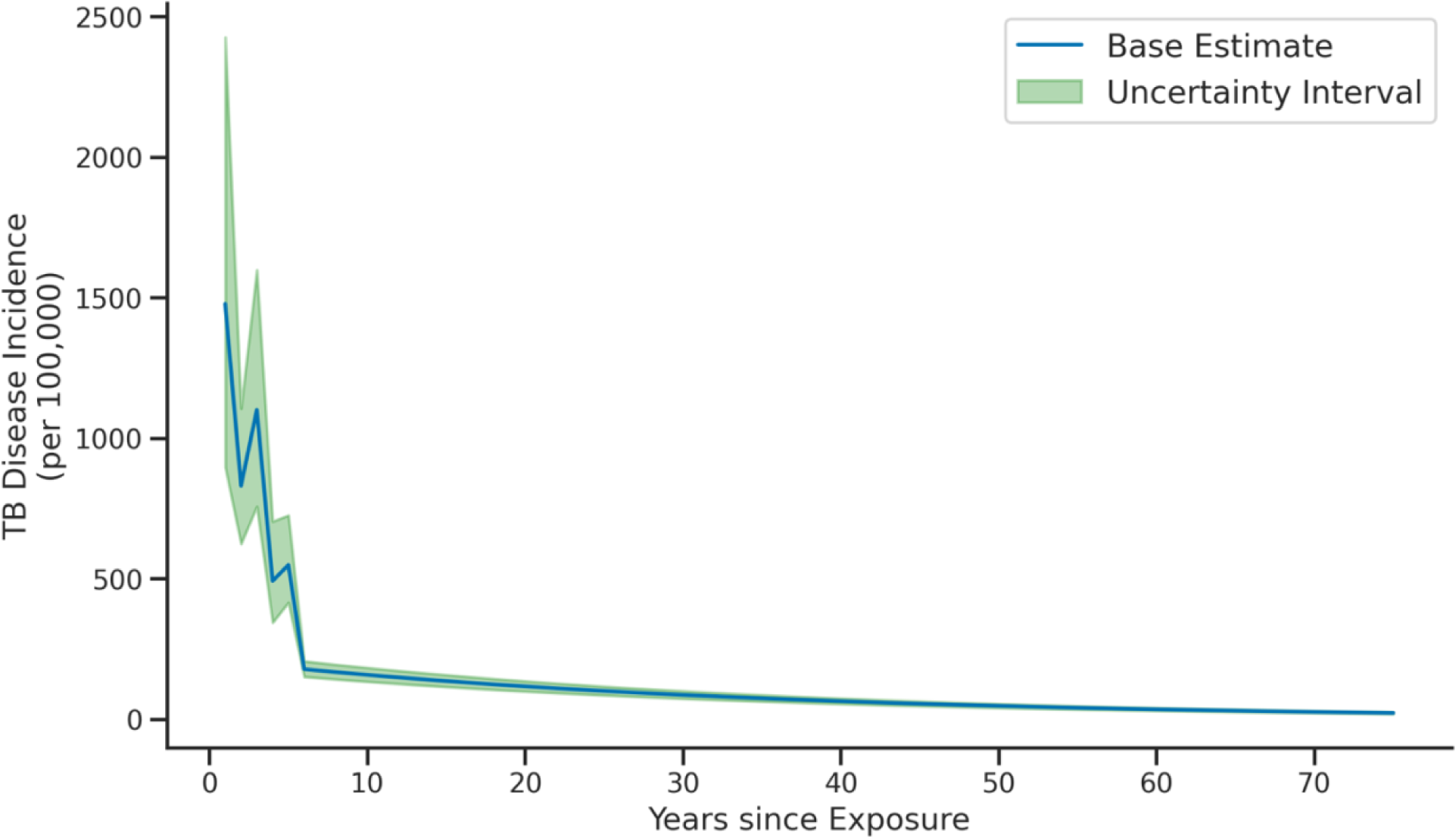
TB incidence assumptions for the model. The first five years, the incidence is elevated as reported in a previous systematic review.(5) Thereafter, we projected that the incidence would drop to the TB incidence rate in India as defined in the WHO Global Tuberculosis Report 2022. We assumed an annual 3% decrease in incidence due to secular reasons.

## Notes

### Competing Interest Statement

The authors have declared no competing interest.

### Funding Statement

This study was supported by the National Institutes of Health (grant number K01AI16773301A1 to PS and a career investment award by the Department of Medicine at Boston University Chobanian & Avedisian School of Medicine to PS Authors were supported by numerous grants during the study which are enlisted in the manuscript. PS also received support from the Warren Alpert Foundation, the Burroughs Wellcome Fund and American Society of Tropical Medicine, and funds from the NIH distributed by CRDF global as well as funds from the Indian department of biotechnology.

### Author Declarations

The study used openly available data from the RATIONS study in India as well as other published data for model parametrization

